# Community-level SARS-CoV-2 Seroprevalence Survey in urban slum dwellers of Buenos Aires City, Argentina: a participatory research

**DOI:** 10.1101/2020.07.14.20153858

**Authors:** Silvana Figar, Vanina Pagotto, Lorena Luna, Julieta Salto, Magdalena Wagner Manslau, Alicia S. Mistchenko, Andrea Gamarnik, Ana María Gómez Saldaño, Fernán González Bernaldo de Quirós

## Abstract

**Background:** By July 1st, the incidence rate of RT-qPCR SARS-CoV-2 infection was 5.9% in Barrio Padre Mugica, one of the largest slums in Buenos Aires City. This study aimed to establish the seroprevalence of SARS-CoV-2 three months after the first case was reported.

**Methods:** Between June 10^th^ and July 1^st^, a cross-sectional design was carried out on people over 14 years old, selected from a probabilistic sample of households. A finger prick sample was tested by ELISA to detect IgG-class antibodies against SARS-CoV-2. Multilevel model was applied to understand sector, household and individual conditions associated with seroconvert.

**Results:** Prevalence based on IgG was 53.4% (95%IC 52.8% to 54.1%). Among the IgG positive cases, 15% reported having compatible symptoms at some point in the past two months. There is evidence of within-household clustering effect (rho=0.52; 95% IC 0.36-0.67); living with a PCR-confirmed case doubled the chance of being SARS-CoV2 IgG positive (OR 2.13; 95% IC 1.17-3.85). The highest risk of infection was found in one of the most deprived areas of the slum, the “Bajo autopista” sector.

**Discussion:** High seroprevalence is shown, for each symptomatic RT-qPCR-confirmed diagnosis, 9 people were IgG positive, indicating a high rate of undetected (probable asymptomatic) infections. Given that transmission among family members is a leading driver of the disease’s spread, it is unsurprising that crowded housing situations in slums are directly associated with higher risk of infection and consequently high seroprevalence levels.

This study contributes to the understanding of population immunity against SARS-CoV2, its relation to living conditions and viral spread, for future decision making.

## Introduction

Population serological data are essential for understanding the prevalence of subclinical infections and the population’s herd immunity against SARS-CoV-2.[1] Several Covid-19 seroprevalence studies were carried out on general population showing, in general, low rate of antibodies.[2-5] However such evidence has been obtained from cities with different living conditions than the ones prevailing among slum dwellers of Latin American cities and can hardly be extrapolated to them. [6]

Buenos Aires City (CABA) has nearly 3 million inhabitants, of whom approximately 5.7% live within 17 neighborhoods with a very vulnerable population. In most cases, there are households with scarce water and sanitation, poor ventilation, space constraints and overcrowding making physical distancing and self-quarantine impractical, thus facilitating the rapid spread of SARS-CoV2 infection.[7][8]

Barrio Mugica is one of the most overcrowded slums with more than 40.000 inhabitants, and almost 1500 people experiencing homelessness. [9] Its first case of SARS-CoV-2 infection, linked to community transmission, was reported four days after national mandatory quarantine was set on March,19^th^ 2020. Preventive measures were taught to community-based organizations that provided food assistance, temporary isolation centers for elderly people in churches were created and an immediate plan for emergency and health care was established in the three medical centers allocated in Barrio Mugica. In spite of these measures, in the following days this slum was the first in Argentina to become affected by a high number of cases, with a sharp rise of infected people.

On May 5th 2020, an active policy named “DETeCTAr”, was set to dampen the spread by testing, diagnosing and early treating of cases.[7] A crisis planning committee joined weekly to enforce the emergency plan actions related to health access, economic aid and food assistance. Social and health community workers were deployed in active surveillance on cases and contact tracing. Local sampling for qPCR testing was performed and each positive person, according to clinical severity, was admitted to an out-of-hospital institution (reconverted hotels) or to a hospital.

Till July 1st, Covid-19 incidence rate at Barrio Mugica was 5.9% (2949 diagnostics by qPCR) and the case fatality rate was 1.5% (44 deaths). In parallel with the mitigation measures the basic reproduction number (R0) in Barrio Mugica decreased from 3.24 to 0.6 in six weeks. In order to understand population immunity against SARS-CoV2, on June, 10th, a team participatory research, the SeroCovid-POB CABA study started, aiming to detect SARS-CoV2IgG presence and the housing conditions associated with infection. The final aim is to contribute to future decision making on epidemiological control measures.

## Materials and Methods

A community-level seroprevalence survey was conducted in a cross-sectional design during a Participatory Action Research (PAR). Population of the study were inhabitants of Barrio Mugica: men and women form 14 years of age or older were included.

Starter PAR team: AG (molecular virologist that contributed to the development of the COVIDAR ELISA Test), ASM (virologist that perform the analysis of the test) and SF (physician that served as volunteer field epidemiologist at the DETeCTAr Barrio 31) offered technical cooperation to test people experiencing homelessness.

PAR-First step: A sample for convenience of 60 homeless people was tested during 3 days in the “Comedor del Fondo”, a popular dining room managed by a non-governmental organization. Samples were collected by a DETeCTAr nurse and a volunteer epidemiologist while training the HCWs. The prevalence was 36%, 22 of them were also RT-qPCR positive and accepted to be inpatient.

PAR-Second step: feasibility was determined by the DETeCTAr coordinators of the Community Health Division of the Ministry of Health and the 14 health community workers (HCWs) when two volunteer epidemiologists from a University Institute explained the field organization (sample technique, data collection and database entry) for obtaining a probabilistic sample of residents. The PAR process was a cornerstone for reaching the sample size.[10][11]

PAR-Third step: A cross-sectional study for seroprevalence survey was carried out.

### Serological test

The test “COVIDAR IgG”, an enzyme linked immunosorbent assay [ELISA] developed and validated in Argentina (Laboratorio Lemos SRL, Buenos Aires, Argentina), was used. Performance characteristics of the kit indicates a sensitivity of 75% after 7 days from symptom onset and 95% after 21 days, using RT-PCR as the gold standard. The specificity was 100% corroborated by studying 200 samples obtained before the pandemic.(data in process of publication) The test detects antibodies against two viral antigens, trimeric spike and the receptor binding domain (RBD) of the spike protein. Viral proteins were expressed in human cells. This kit has obtained regulatory approval by Argentina’s national drug regulatory agency (ANMAT, National Administration for Drugs, Food and Medical Devices)[12]. Blood samples were collected in a capillary tube from a finger prick, taken at the front door of each house.

All HCWs received training for sampling and survey form completion. Epidemiological data were entered in a database. Samples were processed and analyzed at the “Hospital de Niños Dr. R. Gutierrez” Virology laboratory.

### Statistical analysis

Sample size was calculated for a seroprevalence of 6% or higher, accounting for non-response and potential clustering of seropositivity by household (design effect 1.6). A two-stage random sampling method was applied. First level: sector of Barrio Mugica; Second level: geographical areas determined by the Department of Statistic and Census. Thirty houses were selected at this level. People over 14 years old were tested at the front door of their houses.

To obtain the weighted prevalence, the SeroCovid-POB sample dataset was expanded to that of the last census by 3 factors: at neighborhood level, at household level and at individual level. The calculation of expansion factors at the household level is the inverse of the joint probability of selecting the last sampling unit (a household). The expansion factors at the household level imply three types of adjustments. The first one is related with non-response (given that some households did not want to answer the survey); the second one corresponds to the projection of sample to the entire population, and the third one to calibration techniques with a final adjustment by groups of age and gender, using external information from population census.[13] Therefore, calibration variables were people 14 year-old or more, grouped by sex and by intervals of age: 14–30; 31–45; 46-59; 60 and more.

As household members share exposure to SARS-CoV2, the outcome (prevalence of SARS-CoV2 IgG) should show some correlation within the household. To test clustering effect, a random effects logistic regression model (multilevel model) was applied as it includes the variation between clusters explicitly in the likelihood and therefore takes account of intracluster correlation. [14–16] The estimate σ (measure of between households variation) and rho (measure the within-household correlations) were obtained. To test the null hypothesis of no within-household clustering (rho=0) a likelihood ratio test (LRT) was performed comparing the log-likelihood of common logistic regression (in which σ =0 and rho=0) with that for the random effect model.

Explanatory variables were fitted in the models according to a) individual level: age and sex; presence of any related symptom two months before b) household level: cohabiting with a confirmed case, number of households in the building and c) sector level of Mugica: difficulties in water supply and population density. A home ID was set for people living together in a household (defined as those who shared income and food) because in some houses there is more than one household sharing bathroom and kitchen. Associations between explanatory variables and the outcome (positive SARS-CoV2Covid-19 IgG) were determined by estimating the Odd Ratios and their 95% CI from the coefficients obtained in the statistical models. Free license “R software” was used.

The study was approved by the institutional review board of the “Hospital de Niños “Dr R. Gutierrez”. Oral informed consent was obtained from every participant.

The ID number on clinicaltrials.gov public website is NCT04472078.

## Results

The study was carried out between June 10^th^ and June 26^th^, after the curve of incident COVID-19 cases flattened (Figure 1).

**Figure 1:**
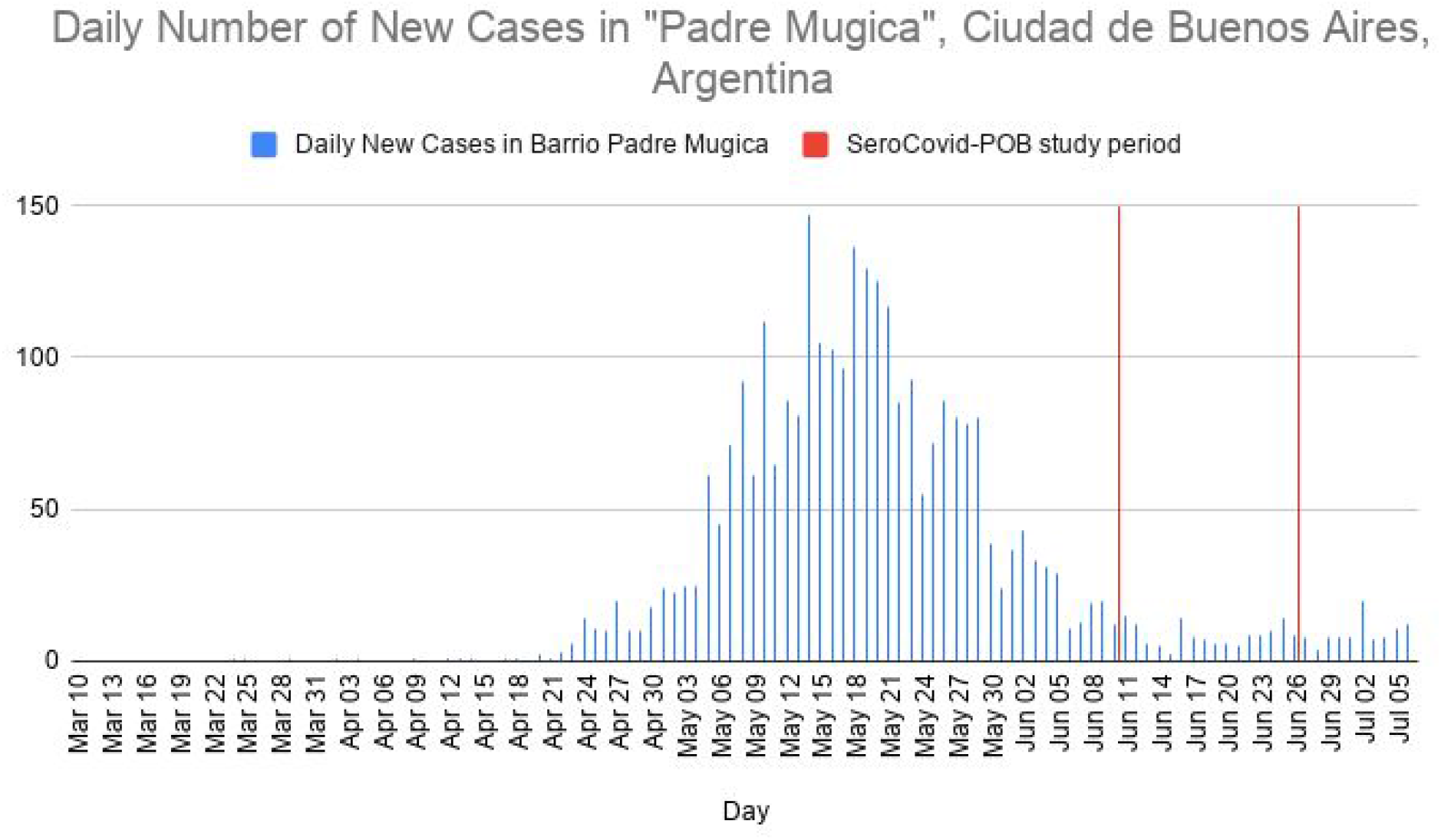
Covid 19 Incident Cases - Barrio Mugica Buenos Aires Argentina. SeroCovid-POB study (June 10th to June 20th).

Barrio Mugica is a heterogeneous slum, in which 10 sectors can be distinguished, each presenting particular cultural and social characteristics.

A total of 398 houses were visited, in which 577 households (families) live. Out of them, 873 people accepted to be tested at their house’s doorstep (Figure 2). Median age was 38 years (IQR 28-49) and 499 (57.2%) were women. Few people (72; 16.8%) reported being symptomatic; 489 houses shared bathroom and kitchen (55.3%). Table 1 shows the descriptive characteristics of the sample according to Barrio Mugica’s sector.

**Table 1:**
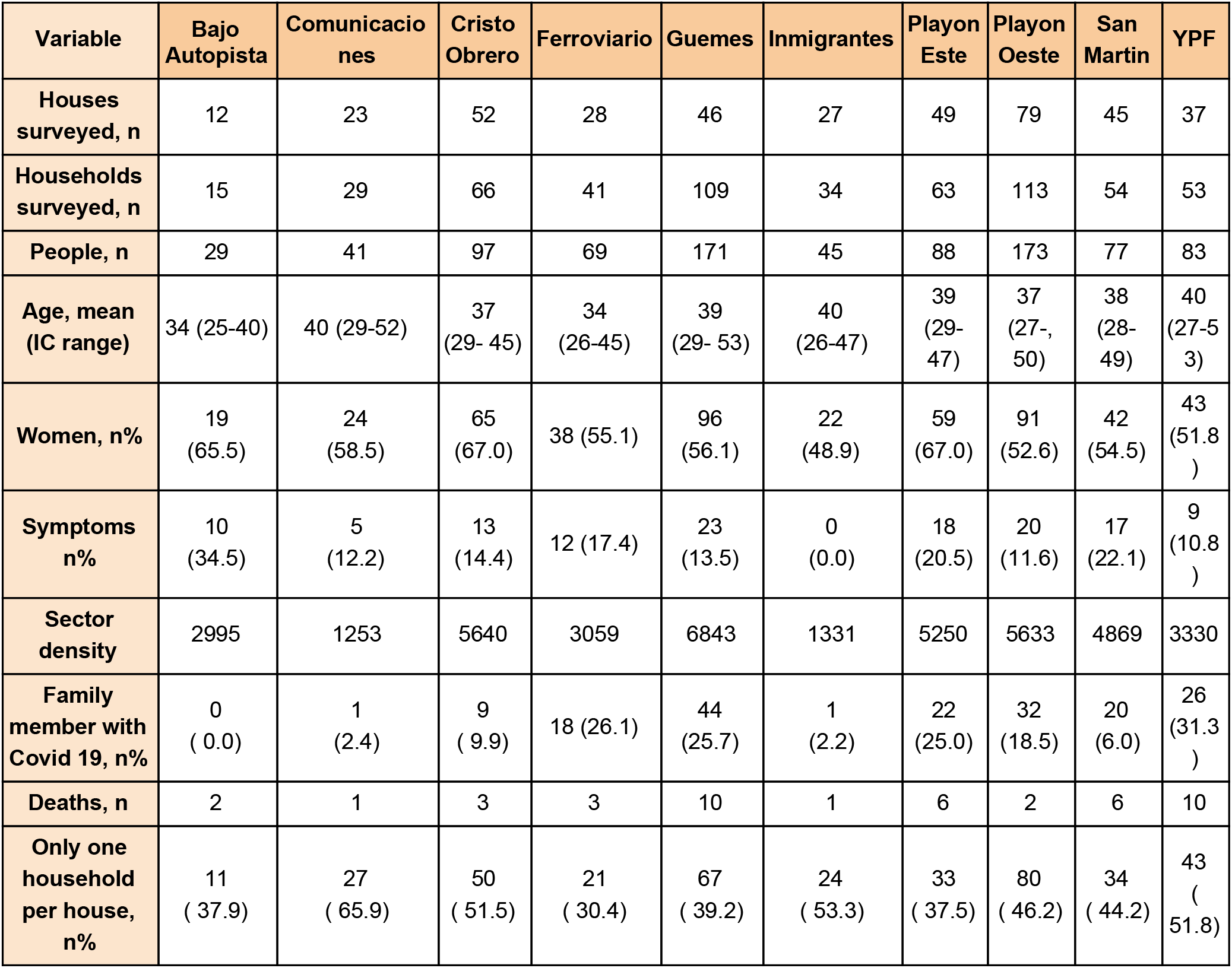
Characteristics of the sample according to Barrio Mugica’s sector of residence.

**Figure 2:**
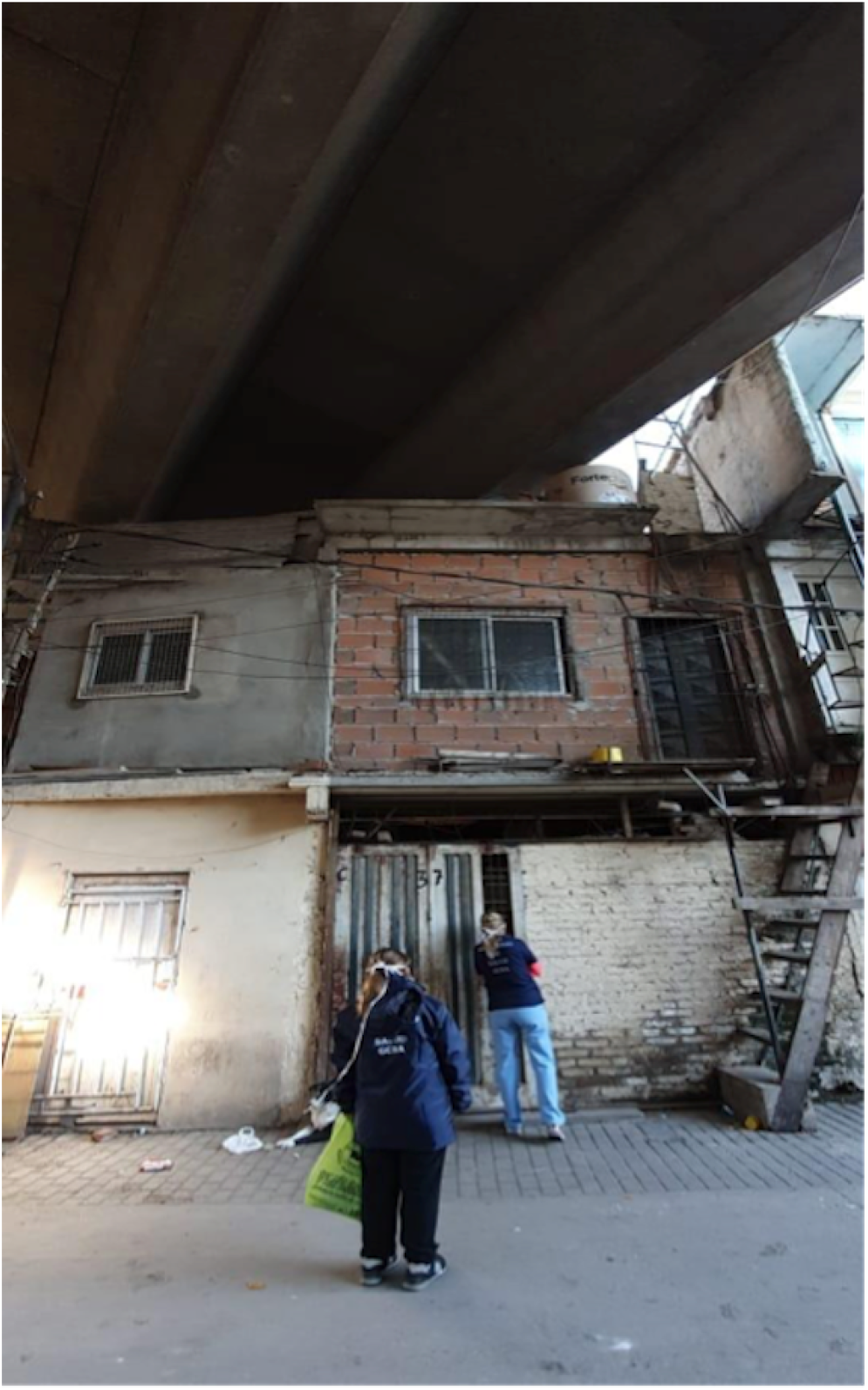
Community Health workers at “Bajo Autopista” sector of Barrio Mugica.

The weighted SARS-CoV2 IgG seroprevalence was 53.4% (95%CI 52.8%-54.1%). Prevalence among males was 51.5% (95%CI 50.6-52.4) and for women 55.3% (95%CI 54.4-56.2). Sex and age-group specific prevalence is shown in Figure 3.

**Figure 3:**
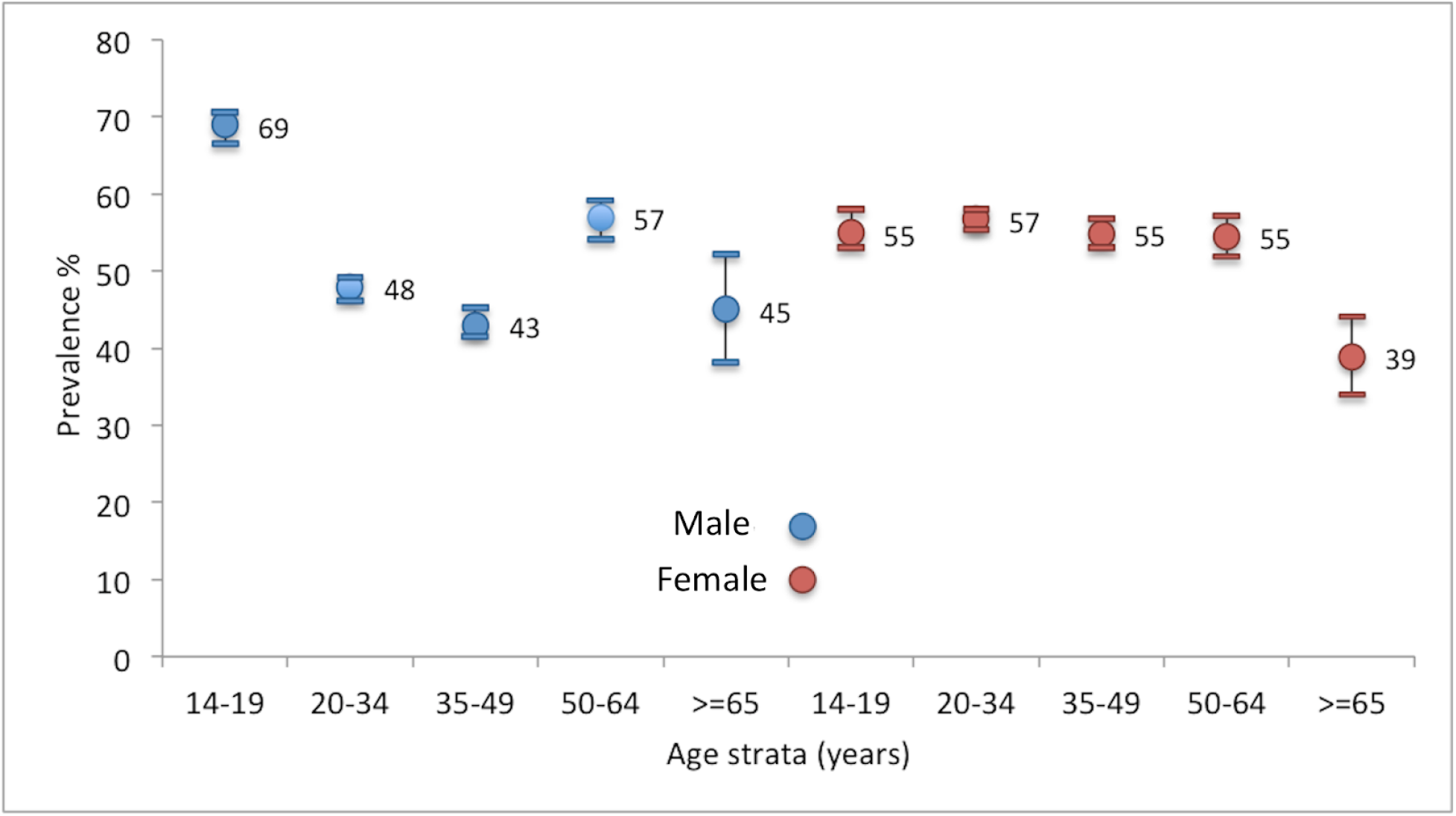
sex and age-strata specific prevalence of positive SARS-CoV2Covid-19 IgG.

Figure shows age-strata specific prevalence by age strata in years with CI95%. Male prevalence in color blue and female prevalence in red.

We have found strong evidence of within-household clustering effect (rho=0.52; 95% IC 0.36-0.67; LRT 43.8 p-value <0.001) and also between-household clustering (sigma 1.88 95% IC 1.37-2.59). We did not find association between being symptomatic and a positive result in serological assay (OR 1.01; 95% IC 0,54-1.87).

The prevalence according to each Barrio Mugica’s sector is shown in Figure 4.

**Figure 4:**
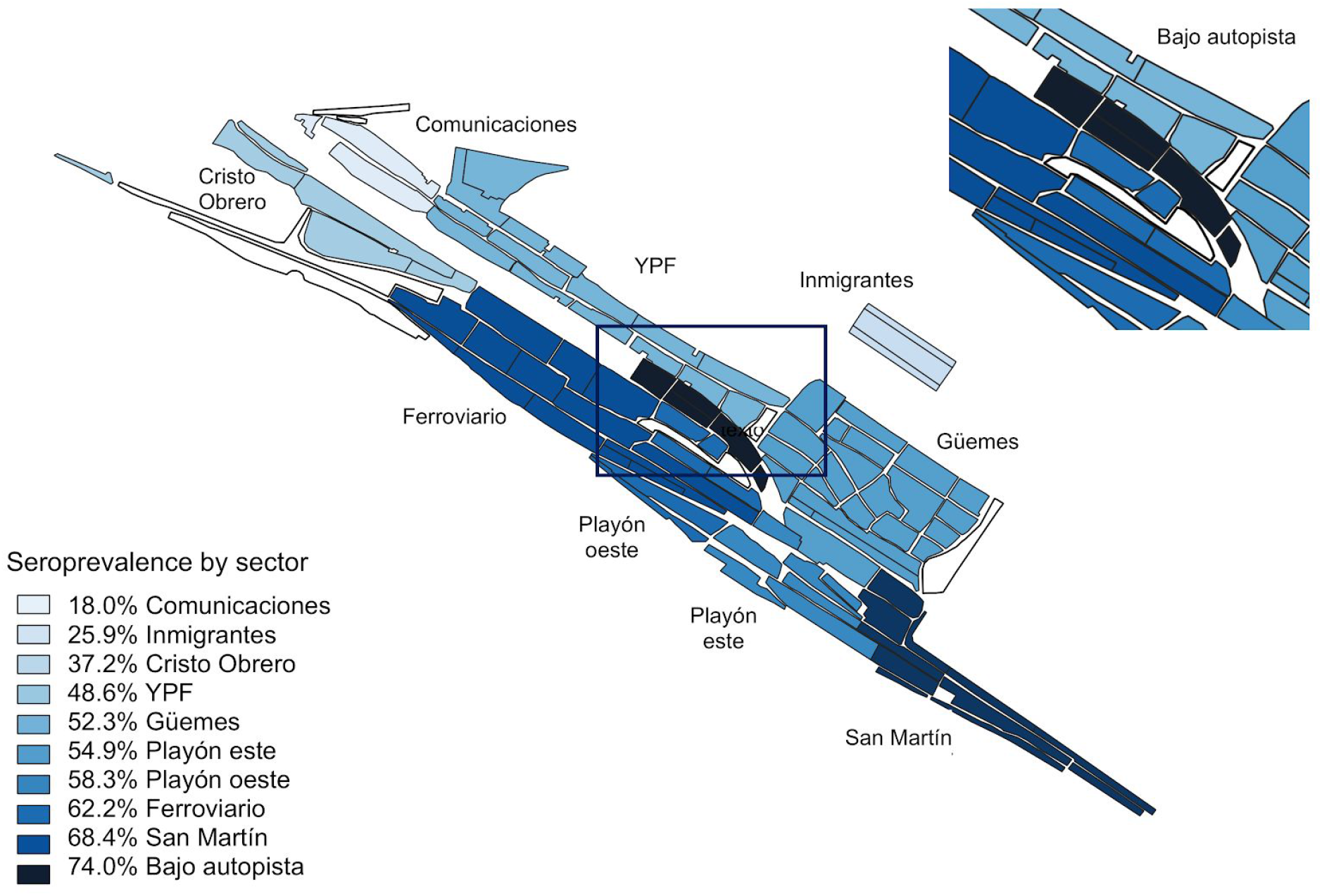
Barrio Mugica Covid-19 seroprevalence by sector.

An association between being IgG positive and living with a PCR confirmed COVID-19 case was found (OR 2.13; 95% IC 1.17-3.85). Living in the “Bajo Autopista” sector of the neighborhood had the highest risk of infection. (OR 30.39 IC95% 5.50-168.0). Other variable associations are shown in Table 2.

**Table 2.**
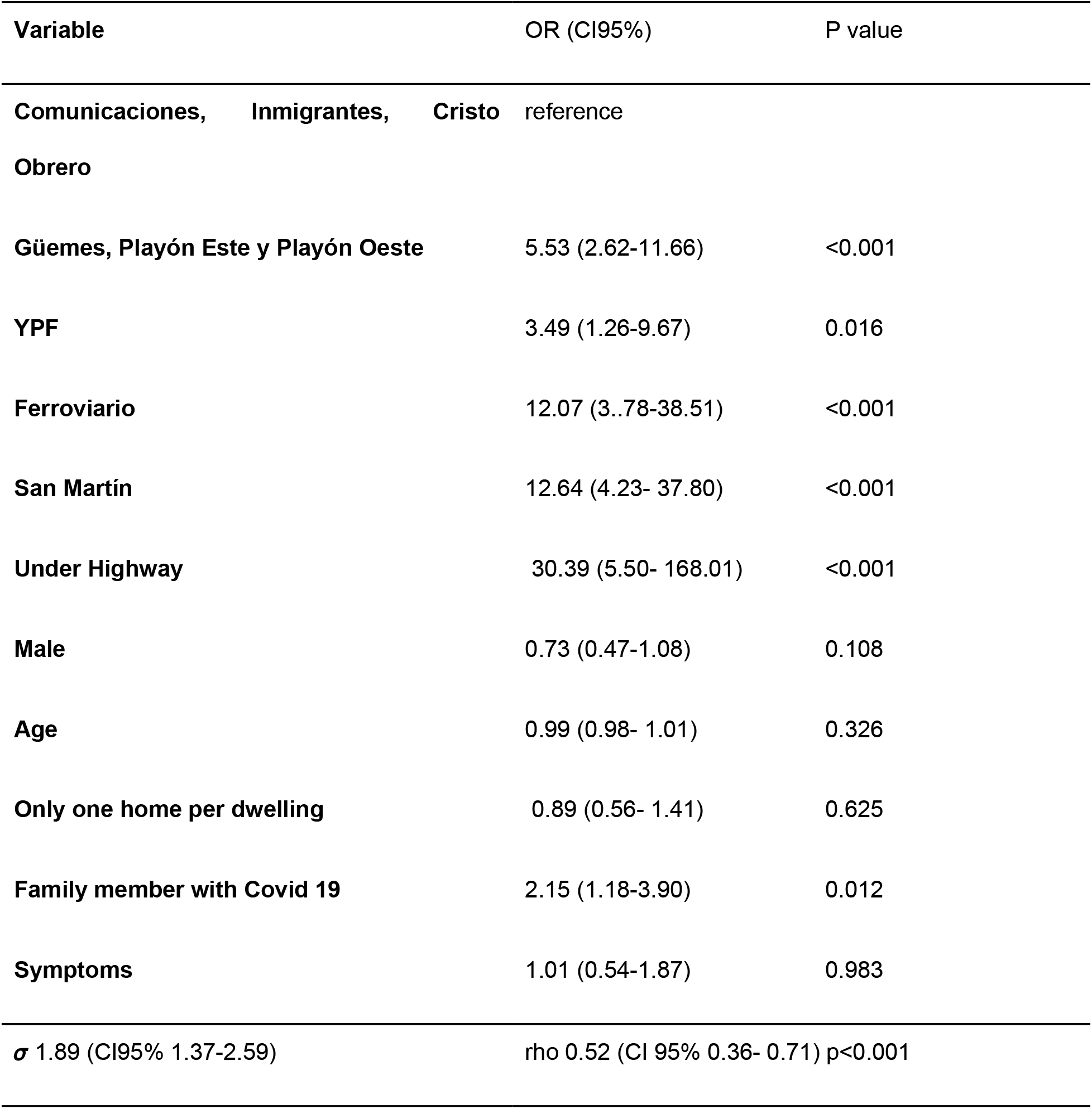
Multivariable model (Random effect logistic regression) to determine whether SARS-CoV 2 IgG is associated with individual, household or sector variables.

## Discussion

Data herein present the first seroprevalence evaluation in the highly vulnerable population in a large slum in Buenos Aires. This city, the capital of Argentina, is a large urban conglomerate in a country of the Southern hemisphere, in which we performed the survey at the beginning of the Winter, a short time after the flattening of the outbreak in the selected slum. The geographical location of the country allowed us to benefit from the previous experiences in the Northern hemisphere, though direct extrapolation was hindered by huge socioeconomic and cultural differences.

The high seroprevalence we found (53%) is in agreement with the recent evidence that COVID-19 has disproportionately affected marginalized populations; the UK and US are reporting higher in-hospital mortality rates for Black people and minority ethnic groups.[17][18] Our study gives an appropriate sample frame tailored to the highly uneven and inequitable distribution of COVID-19 among vulnerable populations.

Our study, a community-level seroprevalence survey, covers a smaller area than large-scale geographic surveys but selects participants in a probabilistic way allowing application to other similar populations. This study helps to understand mechanisms by which SARS-CoV2 spreads in similar low income urban communities. The COVID-19 incidence rate at Barrio Mugica, according to RT-qPCR of suspected cases, was ≥5.9%. Strikingly, we found that in these communities, for each patient diagnosed by RT-qPCR there were 9 IgG-positive individuals, suggesting an unprecedented high rate of asymptomatic infection.

Reporting bias could be present in the symptomatic rate as the survey asked for symptoms during the two previous months, when only people having fever were considered to be COVID-19 cases, thus minimal symptoms may not have been noticed. Even though case definition has changed as new evidence was discovered about the behaviour of the infection around the world, including new mild symptoms, such as lack of taste and smell, this study suggests that in order to avoid major spread of the virus PCR testing criteria should be broader.

One important concern regarding seroprevalence studies that have been previously performed is the specificity of the test used. In this case, an ELISA assay with 100% specific was employed. Regarding sensitivity, the assay showed 95% after 21 days of onset of symptoms using qPCR positive samples. In this respect, if patients included in this study were analyzed before seroconversion, the analysis would be underestimated.

The precise threshold for what percentage of the population would need to be immune against SARS-CoV-2 for herd immunity to occur is currently undefined, however assuming Fine *et al* (2011) projections, if the SARS-CoV-2 basic reproductive number (R0) ranges from 2 to 3.5, this threshold may range from 40% to 75% [20]. In our series, the R0 reached that threshold and the prevalence found is according to this assumption.

A caveat must be kept in mind since sex was not an statistical significant variable, male subjects between 14 and 19 years showed the highest prevalence of IgG positives compared with the other groups.

Due to the high virus transmission found in our study, we would have expected a higher lethality rate. Bonofiglio et al. have shown that in Barrio Mugica 1.5% of people are older than 65 vs 17% in the rest of Buenos Aires City, with a higher working-age population (37% vs 30%), As a working-class neighborhood it has high immigration and elders stay living in their places of origin [9]. In our study, we also found a low proportion of elderly people (4.5%), these differences with the rest of CABA and other european cities could express that, beside a high virus transmission, the lethality rate was not as high as reported in other countries. For future comparison with other countries, we also informed the standardized prevalence, applying WHO population by age and sex was 54.33 (IC95% 53.2 - 55.5).

This study shows the relevance of clustering: cluster correlation was evident among household members who share exposure to COVID-19. In this regard, those who had a family member diagnosed by PCR had two fold chance of becoming infected by the virus. We also found sector differences, those living in worse conditions (“Bajo autopista”, Figure 2) had a higher rate of infection. This could be associated with lack of water supply suffered in April which put the whole community in distressing alert.

The participatory action research process was a cornerstone for reaching the sample size number, because the “community of inquiry and action” evolved, increased and addressed questions that were significant for those who participated in the study. It is important to highlight that co-researchers directly involved in the field work, were community health workers living in the neighborhood [10]. In our research process, people experiencing homelessness were successfully first evaluated, and this encouraged the team to move forward in building capacity to carry out the SeroCovid-POB study. It was important to acknowledge that all stakeholders were part of collective learning relationships. We experienced that PAR challenges the notion that the community is a passive object of study and is devoid of knowledge.

Our study also highlights the challenges that public health officials have to face regarding populations in low-resourced settings, which have not yet been infected. We suggest that top-down strategies to mitigate an infectious disease should incorporate social groups and knowledge that already exist in many slums. Contextual conditions should be recognized in participatory encounters with all the actors working in slum space (foundations, community-based organizations, non-governmental organizations). Community health workers should be trained not only for screening and case and contact tracing but also to challenge the standard ways of treating the urban poor creating more participatory governance.

IgG response against different SARS-CoV-2 antigens becomes detectable in immunocompetent patients after at least 8 days with over 90% of individuals seropositive after day 14 of infection. In theory, seropositive individuals are expected to be at lower risk for re-infection compared to seronegative persons, however neither the level nor the duration of protective immunity against COVID-19 is currently known [17], we will consider this study as a baseline cohort for a serial seroprevalence study that could provide a better understanding of transmission patterns and may help determine when (or if) we reach a state of herd immunity.

## Conclusion

We found an unexpectedly high seroprevalence in a vulnerable population. People living with a PCR confirmed COVID-19 case, increased two-fold the chance of testing IgG positive. Importantly, in this highly populated area with housing deficiencies, for each person diagnosed positive by qPCR with symptoms, 9 people were IgG positive without registering symptoms, providing evidence of high rate asymptomatic infections.

This study directly contributes to understanding population immunity and the housing conditions associated with an unprecedented viral spread, which will help future decision making and control measures.

## Data Availability

All data is available and shown in the manuscript

## Acknowledgement

To the health community workers (HCWs), for their skill, care and joy: Carla Alpire Alponte, Patricia Auza Alarcón, Ayelén Copa Tarqui, Sheila Cortez, Pamela Gallardo, Janeth Gemio Pinaya, Ángeles Hernandez Navarro, Alejandro Maccio, Paula Mosqueda, Nicole Neme, Bania Quispe, Emilio Ramírez Bernal, Thelma Soria, Angélica Fernández Arce. To Susana Guzman who was a cornerstone to train HCWs.

To Mercedes Soriano y Diego Giunta always predisposed to help and encourage us to overcome any research problem.

To all the team of Detectar Barrio 31 to make a friendly context to develop this study in this pandemic chaos.

To COVIDAR Group for providing the ELISA kits and helpful discussions.

To Martin Mendez y Sergio Passamonti for helping us have a deep understanding of cartography.

To Florencia Laborde, for her patience in the revision of the English of the manuscript and help in the submission process.

## Bibliography

[1] Community-level Seroprevalence Surveys | CDC n.d. https://www.cdc.gov/coronavirus/2019-ncov/cases-updates/community-level-seroprevalence-surveys.html (accessed July 4, 2020).

[2] To KK-W, Cheng VC-C, Cai J-P, Chan K-H, Chen L-L, Wong L-H, et al. Seroprevalence of SARS-CoV-2 in Hong Kong and in residents evacuated from Hubei province, China: a multicohort study. The Lancet Microbe 2020;1:e111–8. https://doi.org/10.1016/s2666-5247(20)30053-7.

[3] Stringhini S, Wisniak A, Piumatti G, Azman AS, Lauer SA, Baysson H, et al. Seroprevalence of anti-SARS-CoV-2 ıgG antibodies in Geneva, Switzerland (SEROCoV-POP): a population-based study. Lancet 2020;6736:1–7. https://doi.org/10.1016/s0140-6736(20)31304-0.

[4] Sood N, Simon P, Ebner P, Eichner D, Reynolds J, Bendavid E, et al. Seroprevalence of SARS-CoV-2-Specific Antibodies among Adults in Los Angeles County, California, on April 10-11, 2020. JAMA - J Am Med Assoc 2020. https://doi.org/10.1001/jama.2020.8279.

[5] Pollán M, Pérez-Gómez B, Pastor-Barriuso R, Oteo J, Hernán MA, Pérez-Olmeda M, et al. Prevalence of SARS-CoV-2 in Spain (ENE-COVID): a nationwide, population-based seroepidemiological study. Lancet 2020;0. https://doi.org/10.1016/S0140-6736(20)31483-5.

[6] Pereira RJ, Nascimento GNL d., Gratão LHA, Pimenta RS. The risk of COVID-19 transmission in favelas and slums in Brazil. Public Health 2020;183:42–3. https://doi.org/10.1016/j.puhe.2020.04.042.

[7] Corburn J, Vlahov D, Mberu B, Riley L, Caiaffa WT, Rashid SF, et al. Slum Health: Arresting COVID-19 and Improving Well-Being in Urban Informal Settlements. J Urban Health 2020;97:348–57. https://doi.org/10.1007/s11524-020-00438-6.

[8] Buckley RM. Targeting the World’s Slums as Fat Tails in the Distribution of COVID-19 Cases. J Urban Heal 2020. https://doi.org/10.1007/s11524-020-00450-w.

[9] Bonfiglio JI, Marquez A. Estudios sobre los procesos de integración social y urbana en tres villas porteñas 2017:1–222.

[10] Loewenson R, Laurell AC, Hogstedt C, Ambruoso LD’, Shroff Z. with Alliance for Health Policy and Systems Research (AHPSR) and International Development Research Centre (IDRC) Canada and was launched at the Third Global Symposium on Health Systems Research in South Africa. 2014.

[11] Fuks SI. Systemic Facilitation of Collective Processes: Supporting Creativity and Participative Processes in Groups, Communities and Networks*. 2009.

[12] Reactivos COVID-19 | Argentina.gob.ar n.d. https://www.argentina.gob.ar/noticias/reactivos-covid-19 (accessed July 7, 2020).

[13] Censo de hogares y población: villas 31 y 31 bis, Ciudad de Buenos Aires 2009 (PDF) | Estadística y Censos n.d. https://www.estadisticaciudad.gob.ar/eyc/?p=39240 (accessed July 7, 2020).

[14] Moen EL, Fricano-Kugler CJ, Luikart BW, O’Malley AJ. Analyzing Clustered Data: Why and How to Account for Multiple Observations Nested within a Study Participant? PLoS One 2016;11:e0146721. https://doi.org/10.1371/journal.pone.0146721.

[15] Galbraith S, Daniel JA, Vissel B. A study of clustered data and approaches to its analysis. J Neurosci 2010;30:10601–8. https://doi.org/10.1523/JNEUROSCI.0362-10.2010.

[16] Aarts E, Verhage M, Veenvliet J V., Dolan C V., Van Der Sluis S. A solution to dependency: Using multilevel analysis to accommodate nested data. Nat Neurosci 2014;17:491–6. https://doi.org/10.1038/nn.3648.

[17] COVID-19 deaths analyzed by race and ethnicity — APM Research Lab n.d. https://www.apmresearchlab.org/covid/deaths-by-race (accessed July 7, 2020).

[18] Zhang CH, Schwartz GG. Spatial Disparities in Coronavirus Incidence and Mortality in the United States: An Ecological Analysis as of May 2020. J Rural Health 2020;36. https://doi.org/10.1111/jrh.12476.

[19] Theel ES, Slev P, Wheeler S, Couturier MR, Wong SJ, Kadkhoda K. The Role of Antibody Testing for SARS-CoV-2: Is There One? J Clin Microbiol 2020. https://doi.org/10.1128/JCM.00797-20.

[20] Fine P, Eames K, Heymann DL. “Herd Immunity”: A Rough Guide. Clin Infect Dis 2011; 52:911–916, doi: 10.1093/cid/cir007 https://academic.oup.com/cid/article/52/7/911/299077 (accessed July 7, 2020).

